# CONTRAST SENSITIVITY IS IMPAIRED IN SUSPECTED PRIMARY OPEN-ANGLE GLAUCOMA PATIENTS

**DOI:** 10.1101/2024.03.27.24304979

**Authors:** María Constanza Tripolone, Luis Alberto Issolio, Daniel Perez, Pablo Alejandro Barrionuevo

## Abstract

**Purpose:** To assess contrast sensitivity (CS) for detecting visual changes in suspected POAG patients.

**Methods:** CS was measured foveally at photopic conditions and peripherally at mesopic conditions using sinusoidal gratings of 4 cycles/degree. In experiment 1, foveal and peripheral CS were assessed in suspected POAG patients and age-matched healthy control subjects. In experiment 2, foveal CS was assessed in early POAG patients age-matched with suspected POAG group. Analysis was done considering two age ranges (Under and Over 50 years of age). Correlations between CS and clinical parameters were evaluated.

**Results:** Peripheral CS was decreased only for older POAG suspect patients from the control group (Over 50: p = 0.008. Under 50: p = 0.566). Foveal CS was reduced in POAG suspect participants for both age ranges (Over 50: p = 0.028. Under 50: p < 0.001) and in early POAG patients (Under 50: p = 0.001; Over 50: p < 0.001), both compared to the control group. Foveal CS was lower in early POAG compared to POAG suspect for older patients (Over 50: p = 0.019. Under 50: p = 0.824). Foveal CS was correlated with cup-disc ratio in early POAG patients (Early: p < 0.001. Suspect: p = 0.766) and with age in both patient groups (Early: p = 0.001. Suspect: p = 0.002).

**Conclusion:** CS is affected in patients with a high risk of developing POAG and recently diagnosed. Our results suggest that CS could serve as a screening tool, detecting early damage even before structural changes occur.

## 1 INTRODUCTION

Primary open-angle glaucoma (POAG) is an optic neuropathy characterized by progressive damage of the retinal ganglion cells and the optic nerve, leading to irreversible visual deterioration, and in some cases, to blindness (Mowatt et al., 2008). An early diagnosis, together with adequate treatment, can prevent the progression of the disease, and consequently, improve the patient’s quality of life. Since POAG is a leading worldwide cause of blindness (Zhang et al., 2021), it is essential to explore and identify the changes produced in the very early stage, before an irreparable loss of vision occurs.

There is no precise standard method for detecting POAG at its earlier stages since patients generally do not present other symptoms than a gradual visual field loss (Mowatt et al., 2008; Tatham et al., 2014; Weinreb et al., 2014). Elevated intraocular pressure (IOP), older age, and a family history of glaucoma are some of the risk factors that indicate an increased likelihood of developing POAG (Gedde, Lind, et al., 2021; Gedde, Vinod, et al., 2021). Diagnostic testing for POAG includes structural and functional examination. Evaluation of the optic disc and retinal nerve fiber layer (RNFL) has improved with the development of imaging techniques, such as Optical Coherence Tomography, allowing an objective structural assessment (Hood, 2015, 2017). However, variability in healthy individuals makes it difficult to identify early damage caused by POAG (Weinreb et al., 2014). Standard Automated Perimetry has been a gold standard for evaluating functional deficits. Indeed, visual field scores have been used for categorizing the stage of glaucoma. However, between 30% to 50% of ganglion cells are damaged until a significant loss of visual field is detected (Harwerth et al., 1999; Harwerth & Quigley, 2006; Kerrigan-Baumrind et al., 2000; Quigley et al., 1989). Therefore, visual field tests are useful in more advanced stages of POAG, but not to detect early functional damage.

Contrast Sensitivity (CS) has been measured to evaluate functional vision loss in glaucoma patients (Ichhpujani et al., 2020). Previous studies have found a decreased CS in POAG patients measured at the fovea (Amanullah et al., 2017; Ansari et al., 2002; Bambo et al., 2016; Bierings et al., 2018, 2019; Falcão-Reis et al., 1990; Lahav et al., 2011; Thakur et al., 2018). Furthermore, they reported a significant correlation between CS and cup-disc ratio (CDR) (Lahav et al., 2011), retinal nerve fiber layer thickness (Amanullah et al., 2017), Ganglion Cell/Inner Plexiform Layer Sector Thickness (Fatehi et al., 2017), and mean deviation (MD) visual field score (Bambo et al., 2016; Fatehi et al., 2017; Lahav et al., 2011). Since visual field loss often starts in the periphery, CS was also measured outside the fovea. Peripheral CS sensitivity was found reduced in POAG patients with early visual field loss (Ansari et al., 2002; McKendrick et al., 2007; Thakur et al., 2018). Moreover, in ocular hypertension (OHT) patients peripheral CS was reduced but not so at the fovea (Falcão-Reis et al., 1990). Although there is broad evidence that CS is affected in moderate and advanced POAG, there is not enough information on earlier stages.

Especially important is to investigate if functional changes could provide information of early deterioration caused by POAG. This study aimed to assess CS in patients with suspected POAG. For this purpose, two experiments were carried out. In a first experiment, CS was measured to evaluate differences between POAG-suspect patients and healthy participants in different retinal positions and light levels. In a second experiment, we studied how informative is CS at our best-discrimination condition (obtained from Experiment 1) in relation to results of patients with early POAG. Foveal CS in photopic light conditions was measured in early POAG patients and compared with the results of POAG-suspect participants. Differences between groups were assessed considering two age ranges (Under and Over 50 years of age). Correlations between CS and clinical parameters also were evaluated in early and suspected POAG patients.

## 2 MATERIALS AND METHODS

### 2.1 General methods

#### Participants

Patients were recruited from the Ophthalmology Department of the National University of Tucumán. All patients underwent a complete eye examination including visual acuity (VA), IOP, slit-lamp biomicroscopy, retinography, and visual field test (Octopus 300; Program G1/TOP). The study protocols were approved by the ethics committee of the Universidad Nacional de Tucumán, and were in accordance with the Declaration of Helsinki.

#### Apparatus

A computer-based CS measurement system was used, previously described by Colombo and colleagues (Colombo E. et al., 2009). This system generates visual stimuli that were presented on a widescreen CRT monitor of 17’’ modified with an attenuator to obtain a resolution of 13 bits, about 3500 grey levels after gamma correction (Pelli & Zhang, 1991). The stimuli consisted of achromatic sinusoidal gratings with Michelson contrast between 0,002 and 1. The system allowed the selection of the stimulus spatial frequency within a wide range (1, 2, 4, 8,12, and 24 cycles/degree). Each stimulus was presented at its nominal contrast during 500 ms and was temporally modulated with ascending and descending ramps of 250 ms, making the stimulus appear gradually. Also, it was spatially modulated by a Gaussian function, generating a Gabor patch.

The observer’s eye was placed 1.5 m from the monitor screen, resulting in a visual stimulus size of 4°. Each patch was presented randomly with an inclination of 7° concerning the horizontal clockwise, or counterclockwise. The system employs a discrimination task with a forced choice selection of two alternatives. Using a joystick with two buttons (one for each inclination), the observer must indicate the stimulus orientation, being forced to pick one of them. A modified adaptive psychophysical method based on the QUEST algorithm was used to determine the contrast threshold (Watson & Pelli, 1983). This algorithm makes a Bayesian inference of each response to establish the contrasts of the following stimulus to be presented. Between 25 and 42 trials were required to determine the CS at a specific spatial frequency (the number of trials depends on the observer’s responses)

The system includes normal foveal CS values at the photopic light level obtained on healthy subjects. These normal values were characterized by two age ranges, one from up to 49 years old (Under 50 Years Old) and the second above 50 years old (Over 50 Years Old). For our purposes, we used the normal values, published by Santillán and colleagues (Santillán et al., 2014), to establish the foveal control groups for each age range.

#### Measurement conditions

Foveal and peripheral CS were measured at a spatial frequency of 4 cycles/degree (c/d). This spatial frequency was selected from a previous work (Tripolone et al., 2018), in which at photopic level foveal CS was determined for spatial frequencies of 4 c/d and 8 c/d while at mesopic level foveal and extrafoveal CS was determined for 4 c/d and 2 c/d, respectively. Therefore, 4 c/d was found to be more adequate for evaluating photopic and mesopic CS than 8 c/d and 2 c/d, respectively. For the foveal condition, the measurements were carried out at the photopic level with a stimulus mean luminance of 70 cd/m^2^, favoring the detection of this retinal region. For the peripheral condition, the stimuli were located on the inferonasal retina quadrant. This quadrant was chosen considering that ganglion cell density is higher in the nasal retina than in other regions (Curcio & Allen, 1990; Rodieck, 1998), and the inferior retina presents more vulnerability for glaucomatous damage (Hood, 2017; Hood & Kardon, 2007). A gaze fixation point (red LED) was located on the opposite side of the eye test, at 9 degrees of eccentricity. Because rod density is higher in this retina area than in the fovea (Curcio et al., 1990), the measurements were carried out at the mesopic level. For this condition, using a neutral filter placed just in front of the screen monitor, the stimulus mean luminance was 0.5 cd/m^2^.

The test was conducted monocularly, covering the no-tested eye with an eye patch. If it was possible, both eyes were tested. All participants were dark-adapted for 5 minutes before the session began. For the mesopic range, the participants were also 3 minutes adapted to the mesopic light level. A session required a mean duration of 30 minutes to complete the test in both eyes.

#### Data analysis

The participants were grouped by age ranges (Under 50 Years Old and Over 50 Years Old). Statistical analysis was performed with Minitab software, adopting a significance level of p ≤ 0.05. Differences between groups were determined by Analysis of Variance (ANOVA). Relationships between CS and clinical parameters were analyzed by Pearson correlations.

### 2.2 Specific Methods

#### Experiment 1: Contrast sensitivity in POAG-suspect participants

The diagnosis of suspected POAG was established on clinical findings and/or the presence of risk factors (Gedde, Lind, et al., 2021), based on the following criteria: appearance of the optic nerve or RNFL suspicious for glaucomatous damage without visual field defects, elevated IOP associated with normal appearance of the optic disc, family history of glaucoma. Participants were excluded for any other ophthalmic condition or visual impairment unrelated to POAG.

A total of 34 POAG-suspect participants took part in this study: Age range = 18 – 73 y.o.; VA range = 5/10 – 10/10; Mean Deviation (MD) range = -2.5 – 3.4 dB; IOP range = 12 -25 mmHg; CDR range = 0.3 – 0.8 (asymmetry between both eyes < 0.2).

Healthy control participants, age-matched with the POAG-suspect participants, were recruited from a university cohort. A total of 28 volunteers with normal vision and no ocular or retinal pathology participated to establish the peripheral control group: Age range = 23 – 64 y.o.; VA range = 6/10 – 10/10.

Foveal and Peripheral CS were tested on the POAG-suspect group according to the measurement conditions described in General Methods. Peripheral CS was tested in the healthy control group.

#### Experiment 2: Comparison with early POAG patients

Participants were excluded for any other ophthalmic condition or visual impairment unrelated to POAG. Participants were classified based on visual field results (Mills et al., 2006), and only Early POAG patients were included.

A total of 34 patients were tested: Age range = 20 – 68 y.o.; VA range = 4/10 – 10/10; MD range = -9.54 – 4.4 dB; IOP range = 10 -22 mmHg; CDR range = 0.3 – 0.8. Partial results of this cohort (17 patients) were previously reported (Tripolone et al., 2018). Such as for POAG suspects, Early POAG patients were grouped by age range.

Foveal CS was tested on the Early POAG group according to the measurement conditions described in General Methods.

## 3 RESULTS

A total of 68 participants underwent testing, 34 POAG-suspect patients, 34 early POAG patients, and 28 peripheral control participants. Participants of each group were divided into age ranges: Under and Over 50 Years Old. There was no age-significant difference between POAG-suspect and early POAG groups (*F (1, 66) = 0*.*085; p = 0*.*772*. Under 50 y.o.: *F (1, 17) = 0*.*002; p = 0*.*962*. Over 50 y.o.: *F (1, 47) = 0*.*056; p = 0*.*814*). Between POAG-suspect and peripheral control groups, no age-significant difference was found (*F (1, 60) = 0*.*460; p = 0*.*500*. Under 50 y.o.: *F (1, 18) = 0*.*001; p = 0*.*980*. Over 50 y.o.: *F (1, 40) = 0*.*447; p = 0*.*508*). Foveal control groups were obtained from the system’s normal values. Table 1 summarizes each group data used in our analysis.

**Table 1.**
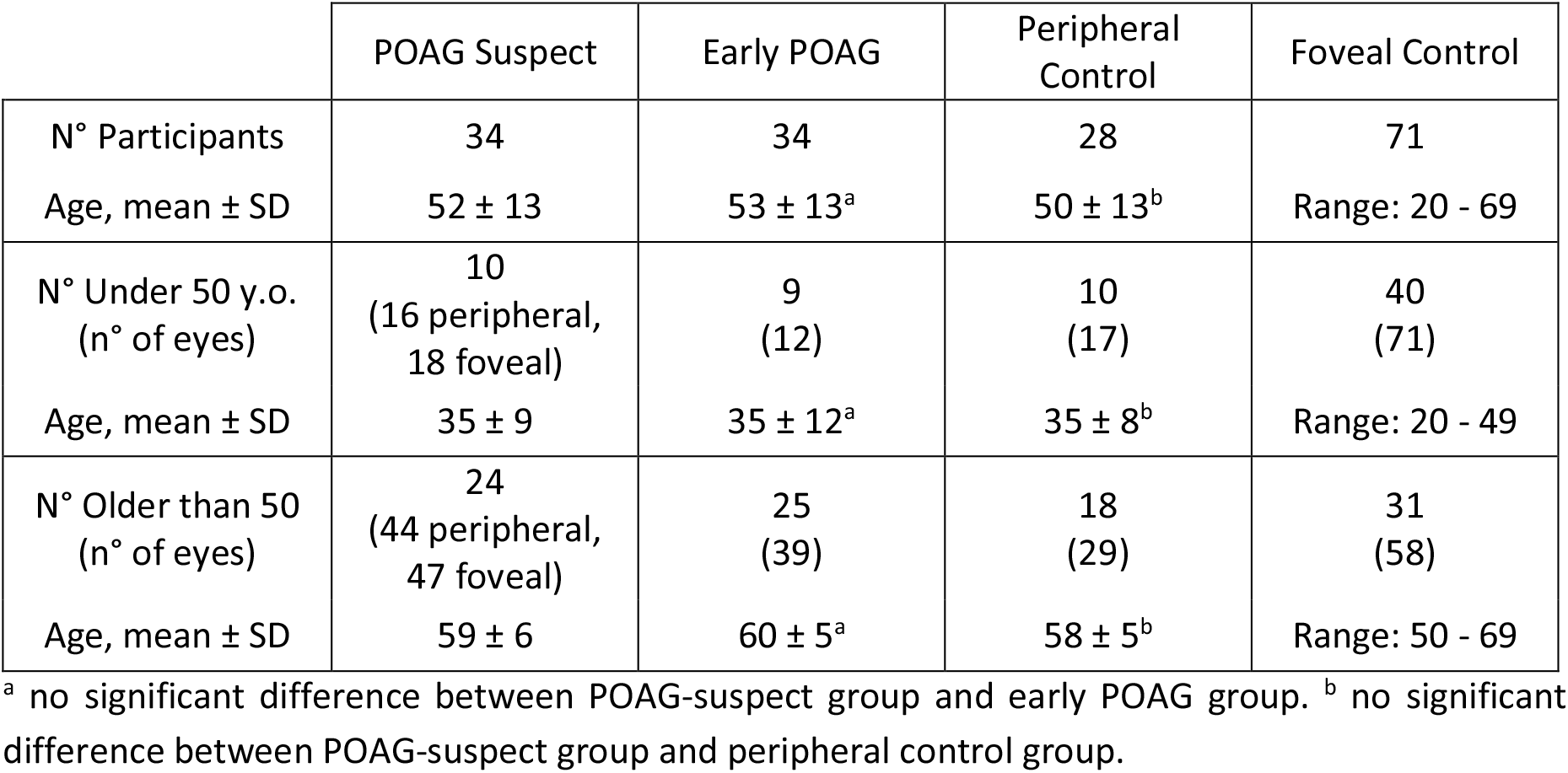
Number of participants for each group.

|  | POAG Suspect | Early POAG | Peripheral Control | Foveal Control |
| --- | --- | --- | --- | --- |
| N° Participants | 34 | 34 | 28 | 71 |
| Age, mean ± SD | 52 ± 13 | 53 ± 13<sup>a</sup> | 50 ± 13<sup>b</sup> | Range: 20 - 69 |
| N° Under 50 y.o.<br>(n° of eyes) | 10<br>(16 peripheral,<br>18 foveal) | 9<br>(12) | 10<br>(17) | 40<br>(71) |
| Age, mean ± SD | 35 ± 9 | 35 ± 12<sup>a</sup> | 35 ± 8<sup>b</sup> | Range: 20 - 49 |
| N° Older than 50<br>(n° of eyes) | 24<br>(44 peripheral,<br>47 foveal) | 25<br>(39) | 18<br>(29) | 31<br>(58) |
| Age, mean ± SD | 59 ± 6 | 60 ± 5<sup>a</sup> | 58 ± 5<sup>b</sup> | Range: 50 - 69 |

<sup>a</sup> no significant difference between POAG-suspect group and early POAG group. <sup>b</sup> no significant difference between POAG-suspect group and peripheral control group.

### 3.1 Experiment 1: Contrast sensitivity in POAG-suspect participants

Figure 1 shows peripheral CS results of POAG-suspect and control groups. Peripheral CS was significantly reduced in POAG-suspect group compared to control group for those over 50 years of age (*F (1, 71)* = 7.38; *p = 0*.*008)*, but not for those under 50 years (*F (1, 31)* = 0.34; *p* = 0.566). The effects of group (POAG-suspect vs control) and age (Under vs Over 50 y.o.) on peripheral CS were not statistically significant (Group: *F (1, 102) = 1*.*68; p = 0*.*198*. Age: *F (1, 102) = 0*.*02; p = 0*.*892*). However, the effect of the group was found to be age-dependent (*F (1, 102) = 4*.*29; p = 0*.*041*). Foveal CS was found significantly decreased in POAG-suspect group compared to control group, for both age ranges (Under 50: *F (1, 87) = 14*.*84; p < 0*.*001*. Over 50: *F (1, 103) = 4*.*98; p = 0*.*028*) (Fig. 2). A significant effect of group (*F (1,90) = 18*.*49; p < 0*.*001*) and age (*F (1, 190) = 26*.*28; p < 0*.*001*) on foveal CS was found. The effect of the group was not age-dependent (*F (1, 190) = 1*.*85; p = 0*.*175*).

**Figure 1.**
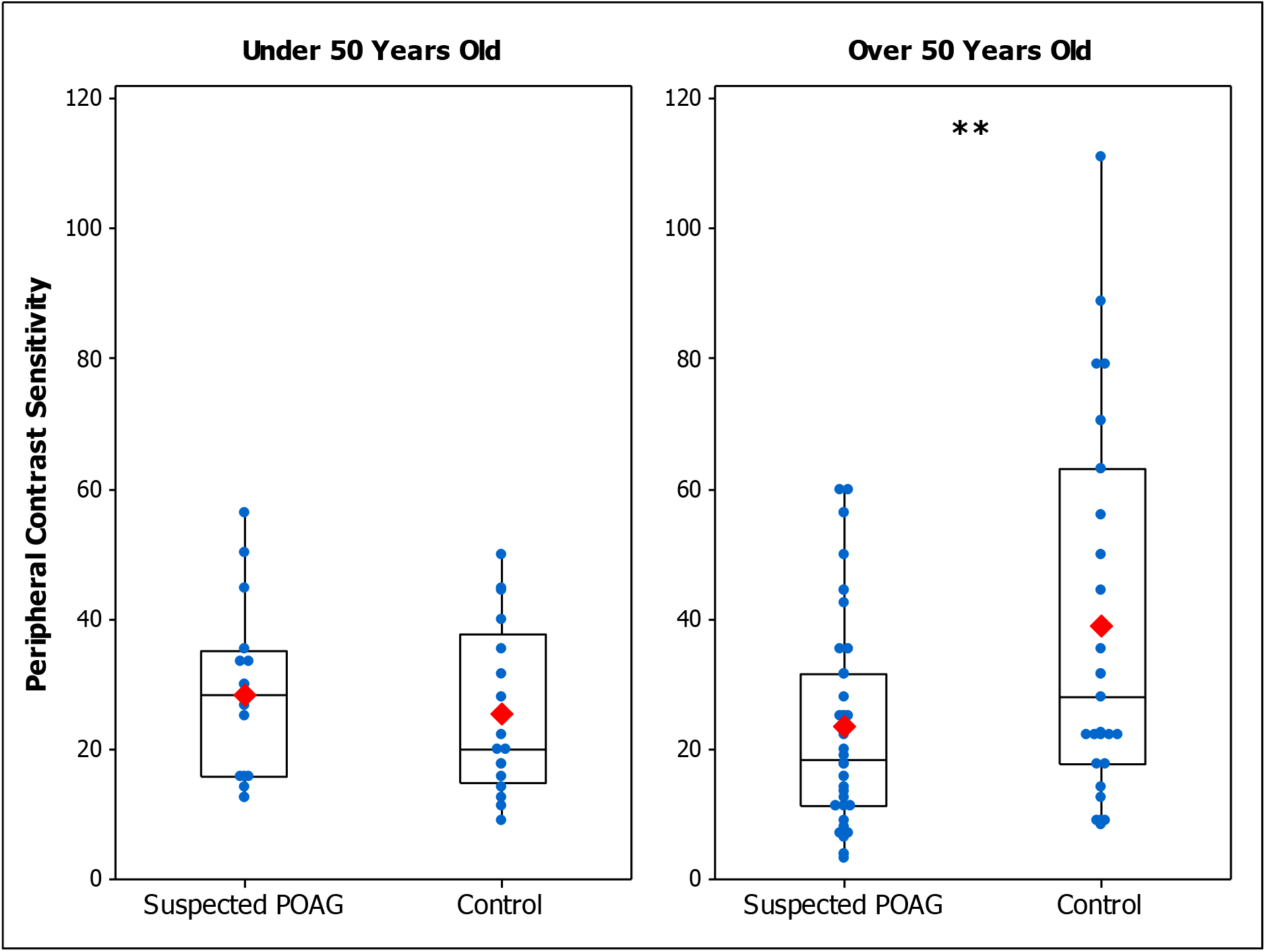
Box plot of peripheral CS in Suspected POAG patients and Control participants for those Under 50 Years Old (left panel) and Over 50 Years Old (right panel). Blue dots are the CS individual’s values and red marks are the mean values of each group. (*) p < 0.05 (**) p < 0.01 (***) p < 0.001

**Figure 2.**
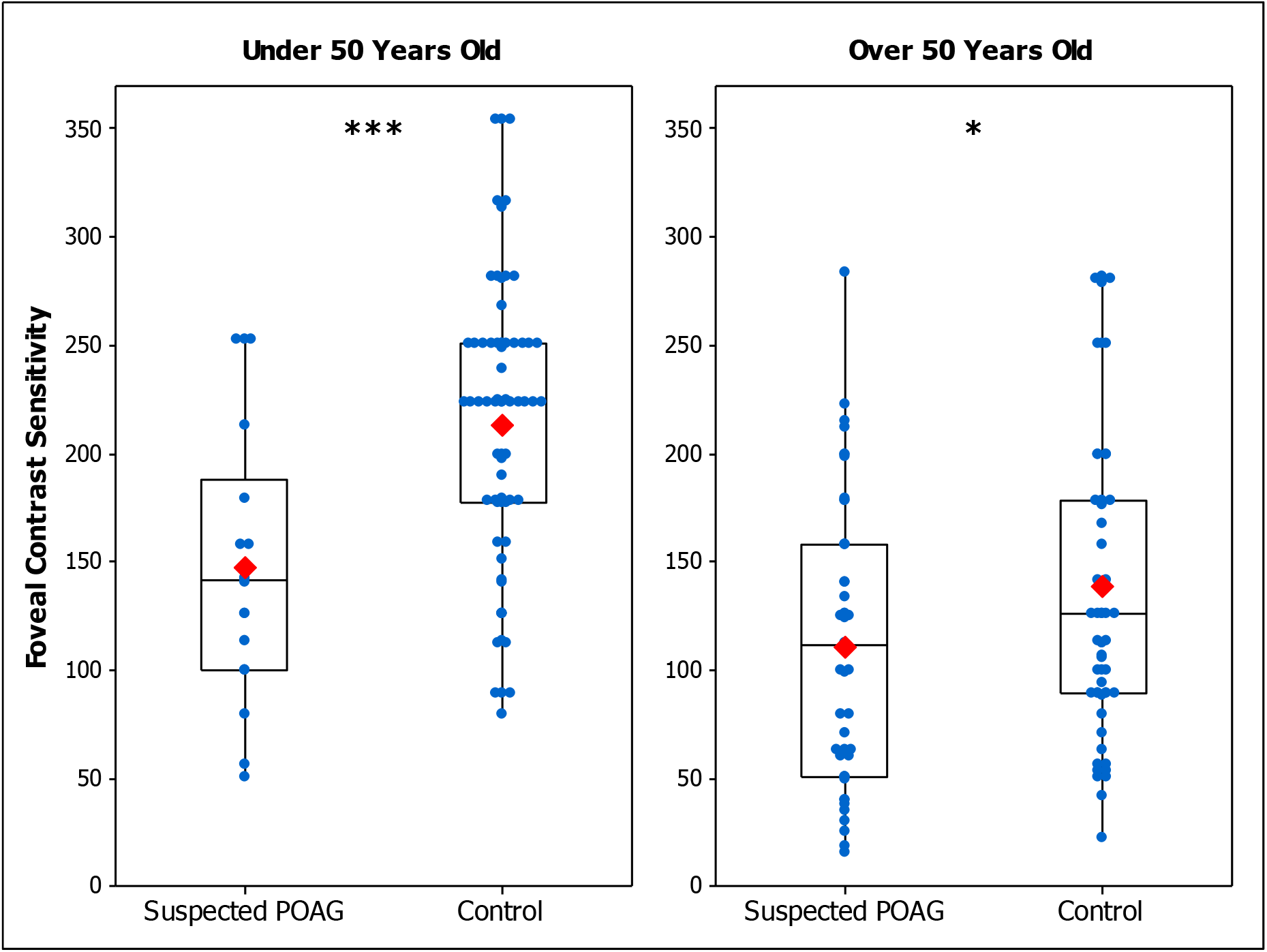
Box plot of foveal CS in Suspected POAG patients and Control participants for those Under 50 Years Old (left panel) and Over 50 Years Old (right panel). Blue dots are the CS individual’s values and red marks are the mean values of each group. (*) p < 0.05 (**) p < 0.01 (***) p < 0.001

The results show a decrease in CS in patients with suspected POAG. Regarding the evaluation in the inferonasal quadrant, CS was found to be reduced only for those over 50 years of age, indicating that the effect of POAG in peripheral CS is age-dependent. On the other hand, the foveal assessment shows a decreased CS in patients for both age ranges, indicating that potential early signs of POAG have an effect on CS and this effect is independent of age. Therefore, foveal results suggest that early visual deficits, before glaucoma diagnosis, could be detected by the CS measurement regardless of the patient’s age.

Since statistical differences between the POAG-suspect group and the control group were found for both age ranges only for foveal conditions, in Experiment 2 we investigated how important the reduction in foveal CS was for POAG-suspect participants in relation with results from patients that were diagnosed as having early POAG.

### 3.2 Experiment 2: Comparison with early POAG patients

#### 3.2.1 Foveal CS in early and suspected POAG groups

Figure 3 shows the foveal CS in early POAG and POAG-suspect patients for each age range. An ANOVA showed that foveal CS in early POAG group is reduced compared to POAG-suspect group for those over 50 years of age (*F (1, 84) = 5*.*72; p = 0*.*019*). However, the difference was not statistically significant for patients under 50 years of age (*F (1, 28) = 0*.*05; p = 0*.*824*). As in our previous study (Tripolone et al., 2018), foveal CS was found reduced in early POAG patients compared to control group for both age ranges (Under 50 y.o.: *F (1, 81) = 11*.*94; p = 0*.*001*; Over 50 y.0.: *F (1, 95) = 21*.*92; p < 0*.*001*).

**Figure 3.**
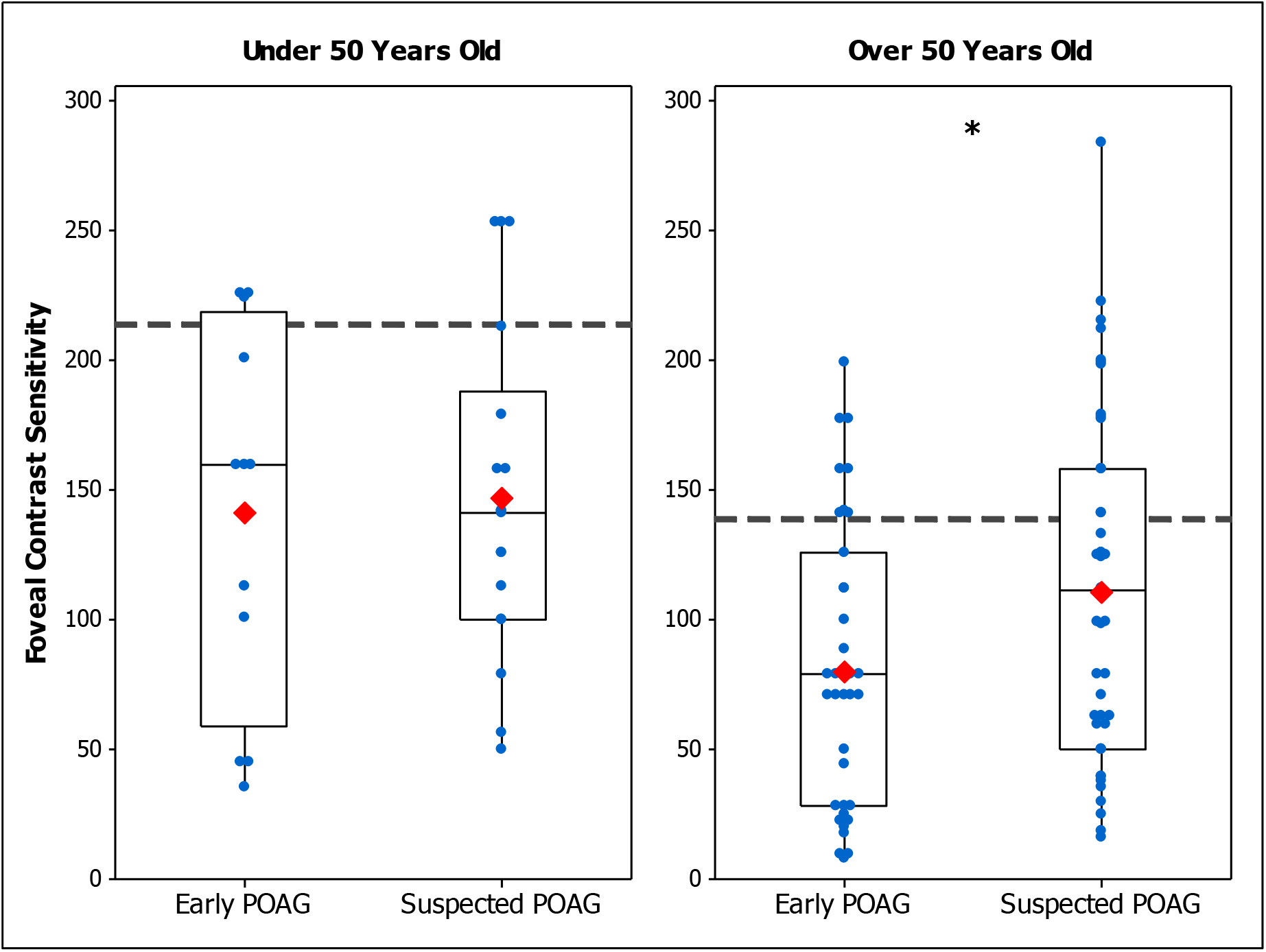
Box plot of foveal CS in Early POAG and Suspected POAG patients Under 50 Years Old (left panel) and Over 50 Years Old (right panel). Blue dots are the CS individual’s values and red marks are the mean values of each group. The mean foveal CS of the foveal control group is shown by the dotted line. (*) p < 0.05 (**) p < 0.01 (***) p < 0.001

These results suggest that foveal CS allows differentiating groups between early POAG from those with suspected POAG when the participants are over 50 years of age. For younger participants, our results show that CS in our POAG-suspect group was as diminished as for early POAG group. In addition, foveal CS measured at the photopic level and 4 cycles/degree can differentiate patients with early and suspected POAG from the control group.

#### 3.2.2 Correlation of Contrast Sensitivity with Clinical Parameters

Pearson correlations were analyzed between CS results and clinical parameters obtained by the eye examination of early and suspected POAG patients. The clinical parameters assessed were: Age, VA, IOP, CDR, and MD. Table 2 summarizes the correlation results.

**Table 2.** Pearson Correlation Coefficients between CS and Clinical Parameters.

|  | Age | VA | IOP | CDR | MD |
| --- | --- | --- | --- | --- | --- |
| Early POAG<br>Foveal CS | $r = -0.45$<br>($p = 0.001$) | $r = 0.21$<br>($p = 0.178$) | $r = 0.24$<br>($p = 0.129$) | $r = -0.53$<br>($p < 0.001$) | $r = 0.09$<br>($p = 0.674$) |
| POAG Suspect<br>Foveal CS | $r = -0.37$<br>($p = 0.002$) | $r = 0.24$<br>($p = 0.076$) | $r = -0.11$<br>($p = 0.446$) | $r = 0.05$<br>($p = 0.766$) | $r = -0.14$<br>($p = 0.527$) |
| POAG Suspect<br>Peripheral CS | $r = -0.21$<br>($p = 0.106$) | not<br>applicable | $r = -0.23$<br>($p = 0.116$) | $r = -0.12$<br>($p = 0.515$) | $r = -0.10$<br>($p = 0.649$) |

$r$ = Pearson Coefficient. $p$ = significance level. The relationship between peripheral CS and VA does not apply due to VA is a foveal measurement.

As expected, foveal CS decreases as age increases (Fig. 4). This association was significant for the three groups (control, suspected POAG and early POAG). In early POAG group was also an indirect significant correlation between CDR and foveal CS, but no correlation was found for POAG-suspect group (Fig. 5).

**Figure 4.**
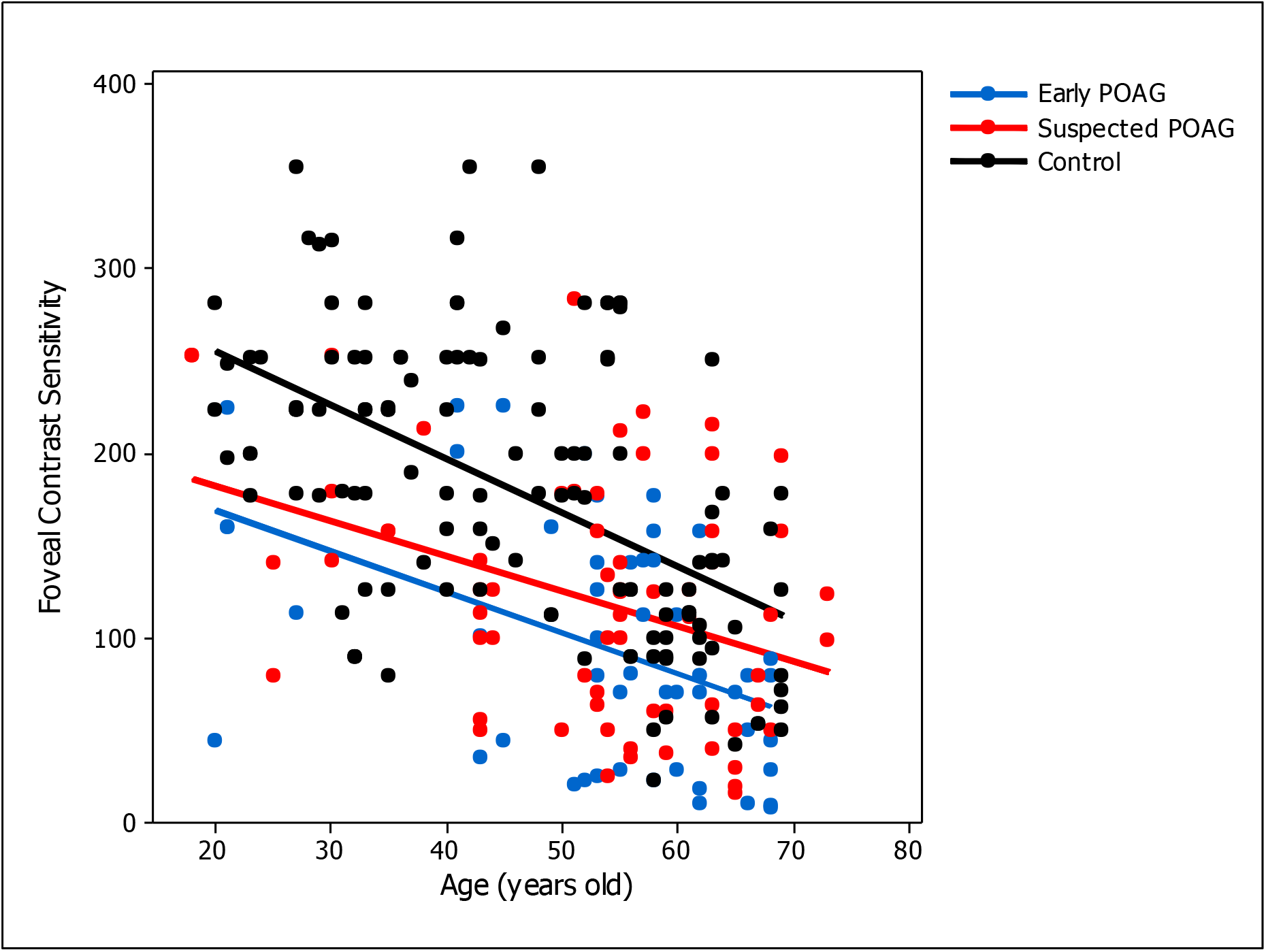
Correlations between foveal CS and age in Early POAG (blue), Suspected POAG patients (red), and Control group (black).

**Figure 5.**
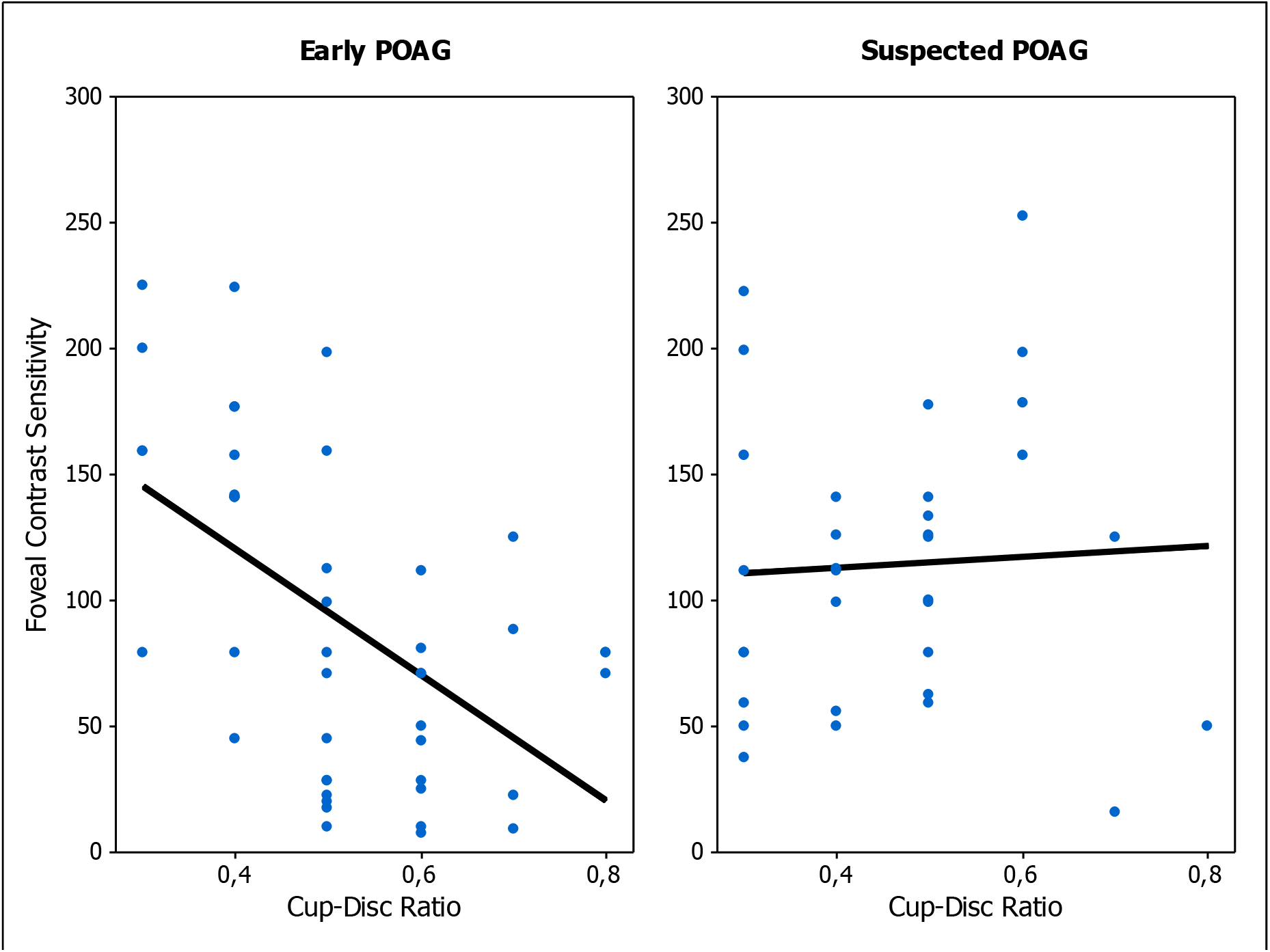
Correlations between CDR and foveal CS in Early POAG (left panel) and POAG-Suspect patients (right panel).

Early POAG group: r = -0.45; p = 0.001. POAG-Suspect group: r = -0.37; p = 0.002. Control group: r = -0.53; p < 0.001. Lines represent the linear model for each patients’ group [Early POAG (blue): slope = -2.21, intercept = 213.45; Suspected POAG (Red): slope = -1.9, intercept = 220.17; Control (black): slope = -2.91, intercept = 314]. Control data was obtained from the study of Santillan and colleagues (Santillán et al., 2014).

Early POAG group: r = -0.53; p < 0.001. POAG-Suspect group: r = -0.06; p = 0.766. Black lines represent the linear model.

## 4 DISCUSSION

CS was assessed in patients with suspected primary open-angle glaucoma in different experimental conditions and in comparison, with early POAG patients. Peripheral assessment showed a decreased CS for patients over 50 years of age, but not for the younger group with respect to healthy age-matched control groups. Foveal CS was reduced in POAG suspect patients compared to healthy age-matched control participants for both age groups (under and over 50 years old). Similar results were found in early POAG patients, with foveal CS reduced for both age groups with respect to the healthy cohort. We found a lower foveal CS for the early POAG group when compared to the POAG suspect group for participants over 50 y.o. However, we didn’t find a difference between these groups for patients under 50 y.o. Together, these results show how contrast sensitivity is affected in patients with high risk and at the very early stage of POAG, and evidence the potential of CS to detect first manifestation of this optic neuropathy.

Peripheral CS was previously found diminished in POAG (Falcão-Reis et al., 1990), early to moderate POAG (Ansari et al., 2002), and early POAG patients (Lundh & Gottvall, 1995; McKendrick et al., 2007), in agreement to our results in POAG suspect subjects. However, in ocular hypertension (OHT) patients CS evaluated at 15° in the temporal retinal quadrant remained the same as the control group (Ansari et al., 2002). The CS impairment found in our study at the infero-nasal quadrant (9° of eccentricity), suggests that this retinal area could be more sensitive to early visual changes. Indeed, the inferior retina represents a more vulnerable zone to be affected in the early stages of glaucoma (Hood, 2017; Hood et al., 2013). In another study involving OHT patients with a high risk of developing POAG, differences with age-matched controls were found at high eccentricities (20° and 25° outside the fovea) but not at lower (10° and 15°) at photopic level (Falcão-Reis et al., 1990). Explanations of these spatial differences might include optical aberrations, light level conditions and photoreceptor signaling, however these speculations are out of the scope of the present study.

Regarding foveal CS, several studies have reported a decreased CS in patients with different stages of glaucoma (Bambo et al., 2016; Eshraghi et al., 2019; Lahav et al., 2011; Onal et al., 2008; Richman et al., 2015; Thakur et al., 2018). However, there is little evidence about CS in previous stages of POAG. Falcão-Reis and colleagues found decreased foveal CS in POAG patients, although high-risk OHT patients did not present differences with the control group (Falcão-Reis et al., 1990), in disagreement with our results in suspected POAG patients. Since they used flickering stimuli while we used static patterns, we can argue that static patterns are more appropriate than flickering ones for detecting CS defects in POAG-suspect patients. Indeed, Ansari and colleagues found a large, but not significant, CS difference between normal and OHT patients with stationary compared to flickering gratings (Ansari et al., 2002). On the other hand, a trend of decreasing photopic CS with increasing severity of glaucoma have been previously shown (Fatehi et al., 2017; Lahav et al., 2011), in agreement with our results in which a lower CS reduction was found in POAG suspect patients compared with early POAG patients.

Considering the spatial frequency, the magnitude of the depression in CS for early POAG patients was reported to be larger at spatial frequencies around the peak contrast sensitivity function than at other spatial frequencies (Bierings et al., 2019; Wood & Lovie-Kitchin, 1992). Since for our experimental conditions a peak CS was found around 4 c/g for healthy observers (Santillán et al., 2014), the assessment at maximum CS could be more appropriate to evaluate early visual changes caused by POAG.

In comparison with other clinical tests, it was shown that the structural impairment measured by the CDR was correlated with foveal CS in early POAG patients, in agreement with previous evidence of significant correlations with ganglion cell macular thickness (Fatehi et al., 2017) and CDR (Lahav et al., 2011). This association suggests that CS decreases as the damage to the optic disc increases and it provides further support of our results. However, in POAG-suspect subjects, neither foveal nor peripheral CS were correlated with CDR. Since POAG-suspect participants were patients with a risk of developing glaucoma, these results suggest an impairment of CS previous to established optic disc damage. Considering functional visual tests, CS was not correlated with VA and MD. Since VA is the minimum size detectable of a high contrast pattern, it is possible that this function is related to CS measured at high than at intermediate spatial frequencies. On the other hand, correlations between foveal CS and MD have been previously reported in early to advanced glaucoma patients (Bambo et al., 2016; Fatehi et al., 2017; Lahav et al., 2011). As our cohort included only patients with no or mild visual field loss, MD was almost not expected to be associated with CS, suggesting that CS impairment precedes visual field scores changes

The effect of participants’ age in CS has been largely studied (Derefeldt et al., 1979; McKendrick et al., 2007; Santillán et al., 2014; Zhuang et al., 2021) demonstrating that older adults have lower CS than younger ones, caused by age changes of optical and neural factors. Indeed, a marked CS difference in healthy individuals was established between groups under and over 50 years of age (Santillán et al., 2014; Zhuang et al., 2021). For this reason, it is essential to differentiate age ranges when comparing patients with healthy individuals. Thus, CS differences between these groups could be attributed to the disease and not to the age of the participants. Our results showed a significant association of foveal CS with age for both early and suspected POAG patients, indicating an effect of age in CS. This effect was similar to the one found for the control group (Fig. 4). Each patient group (early and suspected) were age-matched with the control group, implying that the decreased of CS found in these patients is mainly caused by visual deficits of this optic neuropathy and not because of the participants’ age. Interestingly, foveal CS of POAG suspect patients was more similar to early POAG results for young than for older participants (Figs. 3 and 4), suggesting that CS might be more informative at young age. However, this trend was not found for peripheral conditions, where CS was only reduced in POAG-suspect group for participants over 50 years of age, suggesting that peripheral CS is not informative for a young cohort.

Diagnosing POAG at its ultra-early stages continues to be a challenge. Optical coherence tomography has improved the assessment of early structural damage in suspected POAG patients (Hood, 2017; Stagg & Medeiros, 2020). Assessment of the visual field has been demonstrated to be adequate for diagnosing and assessing of progression of POAG at more advanced stages (Medeiros et al., 2012; Quigley et al., 1989). Functional changes in POAG suspects also were evaluated by chromatic pupillometry (Adhikari et al., 2016) and electroretinography (Banitt et al., 2013; Tirsi et al., 2022), reporting an early deficit in ganglion cell function in those patients. There is wide evidence of reduced CS in POAG patients. Our study provides evidence that such reduction occurs also in POAG-suspect eyes and can be evidenced in the visual function.

Despite CS is a potential measure for exploring the visual system, it has not been broadly implemented in clinical settings, possibly, because faster tests are preferred. Our protocol for measuring peripheral and foveal CS was relatively long duration for assessing both eyes (approximately 30 minutes including adaptation periods). Instead, we propose a reduced protocol, measuring foveal CS (photopic, 4 c/g), lasting about 10 minutes to test both eyes and also being appropriate to examine patients in a wide age range. Considering individual values, there was an overlap between patients and healthy controls (Fig. 1, 2 and 3). Thus, CS could serve as a screening tool, even when the visual field is within normal values and VA is preserved.

Our findings show evidence of the valuable role that CS plays, providing more information than other tests in suspected POAG patients. Therefore, our study demonstrates that retinal functioning is affected before structural damage even in patients with a risk of developing primary open angle glaucoma.

## Data Availability

All data produced in the present study are available upon reasonable request to the authors

## ACKNOWLEDGEMENTS

This work was supported by the Agencia I+D+i [grant code PICT 2019-03673], the Consejo Nacional de Investigaciones Científicas y Técnicas [grant codes PIBAA-1234 and PIP-2721] and the German Research Foundation (DFG) [project number 222641018 - SFB/TRR 135 TPs C2 and B2]. We thank Dr. Javier Santillán for providing the foveal control data.

